# Deep learning NTCP model for late dysphagia after radiotherapy for head and neck cancer patients based on 3D dose, CT and segmentations

**DOI:** 10.1101/2025.06.20.25329926

**Authors:** S.P.M. de Vette, H. Neh, L. van der Hoek, D.C. MacRae, H. Chu, A. Gawryszuk, R.J.H.M. Steenbakkers, P.M.A. van Ooijen, C.D. Fuller, K.A. Hutcheson, J.A. Langendijk, N.M. Sijtsema, L.V. van Dijk

## Abstract

**Background & purpose:** Late radiation-associated dysphagia after head and neck cancer (HNC) significantly impacts patient’s health and quality of life. Conventional normal tissue complication probability (NTCP) models use discrete dose parameters to predict toxicity risk but fail to fully capture the complexity of this side effect. Deep learning (DL) offers potential improvements by incorporating 3D dose data for all anatomical structures involved in swallowing. This study aims to enhance dysphagia prediction with 3D DL NTCP models compared to conventional NTCP models.

**Materials & methods:** A multi-institutional cohort of 1484 HNC patients was used to train and validate a 3D DL model (Residual Network) incorporating 3D dose distributions, organ-at-risk segmentations, and CT scans, with or without patient- or treatment-related data. Predictions of grade ≥2 dysphagia (CTCAEv4) at six months post-treatment were evaluated using area under the curve (AUC) and calibration curves. Results were compared to a conventional NTCP model based on pre-treatment dysphagia, tumour location, and mean dose to swallowing organs. Attention maps highlighting regions of interest for individual patients were assessed.

**Results:** DL models outperformed the conventional NTCP model in both the independent test set (AUC=0.80-0.84 versus 0.76) and external test set (AUC=0.73-0.74 versus 0.63) in AUC and calibration. Attention maps showed a focus on the oral cavity and superior pharyngeal constrictor muscle.

**Conclusion:** DL NTCP models performed better than the conventional NTCP model, suggesting the benefit of using 3D-input over the conventional discrete dose parameters. Attention maps highlighted relevant regions linked to dysphagia, supporting the utility of DL for improved predictions.

## Introduction

Dysphagia is a common late side-effect of radiotherapy for head and neck cancer (HNC), which impairs patients’ quality of life immensely by limiting their intake.^1^ Furthermore, dysphagia may be associated with an increased risk of aspiration and consequently aspiration pneumonia, a potentially life-threatening disease.^2^ Normal tissue complication probability (NTCP) models can estimate the risk of developing dysphagia in HNC patients following radiotherapy and are typically based on discrete organ at risk (OAR) dose values and/or patient characteristics.^3,4^ Radiation-associated dysphagia (RAD) risk estimation can be used to guide dose optimization and treatment decision-making, such as selecting patients for either photon or proton therapy.^5,6^

Swallowing is a complex mechanism, involving over 30 muscles and nerves that collaborate to move ingesta from the mouth to the stomach.^7–9^ Current NTCP models predicting late RAD include dose-volume parameters of pharyngeal constrictor muscles (PCMs)^3,4^, either in combination with the supraglottic larynx^3^, oral cavity^4^ or brainstem^10^ as predictors. However, this is a limited representation of swallowing OARs, since not all OARs related to swallowing are included in these models.^11–13^ Conventional logistic regression NTCP models cannot include dose parameters of all relevant swallowing muscles due to multicollinearity (i.e. high correlation between parameters) and overfitting (i.e. creating a complex model that does not generalise to new patients).^14^ Furthermore, dose parameters in NTCP models are usually mean doses or dose-volume thresholds to specific OARs, which may oversimplify the given dose distribution, losing important spatial details.^13^ Hence, there is a need for more advanced modelling that can deal with complex radiation dose data, extracting information from 3D dose distributions and imaging information of all regions involved in swallowing.

Deep learning (DL) methods have the potential to process complete 3D dose distributions and CT scans to predict a toxicity risk, without reducing radiation dose to single values (1D) like in conventional NTCP models. Furthermore, by utilising comprehensive anatomical information from CT scans and radiation dose distributions, DL models can identify complex relationships between radiation dose and all OARs visible on the CT scan. This represents an additional advantage over conventional logistic regression NTCP models, as it eliminates the need for manual selection of model parameters. Additionally, DL models can better deal with multicollinearity compared to conventional NTCP models, as DL models automatically reduce overlap between related input factors.^15,16^ Moreover, incorporating clinical variables such as tumour location and pre-existing conditions (i.e. baseline dysphagia), can be done in both conventional and DL models to provide all relevant information for the prediction of late toxicities.^2,4,17,18^ For xerostomia (i.e. dry mouth), DL models have already demonstrated superior performance compared to conventional NTCP models and highlight sub-regions of the parotid glands as important structures for xerostomia prediction.^19,20^ This approach using DL may be even more suitable for predicting late RAD, given the interconnected nature of swallowing muscles, the complex damage mechanisms involved, and the variety of data that can be used for predicting RAD (e.g., pre-treatment factors, CT scans, and dose distributions).

Therefore, the aim of this study was to improve the prediction of RAD at six months after treatment with DL compared to a validated conventional NTCP model for HNC patients. Additionally, this study explored multiple methods to incorporate clinical variables in a DL model.

## Methods and materials

### Patient data

Patients were treated at the University Medical Centre Groningen (UMCG) and MD Anderson Cancer Centre (MDACC) between 2007-2021 and 2015-2021, respectively. Patients met the following inclusion criteria: 1) squamous cell carcinoma (originating from the oral cavity, pharynx, or larynx), 2) treated with definitive curative HNC radiotherapy, 3) finished all planned treatment fractions, 4) dysphagia assessment available pre-treatment and 6 months after RT, 5) were 18 years or older, 6) did not have previously treated HNC or surgery in the head and neck region (except for tonsillectomy or laser treatment for small glottic lesions).

The UMCG cohort was used for DL model development (85%: 70% training, 15% internal validation) and testing in an independent never-seen-by-the-model set (15%); this split was stratified by treatment modality (photon-vs proton-treatment), contrast-enhancement of the CT, and CT artefacts. The MDACC cohort was used for external validation.

In the UMCG, patients were treated with 66-70 Gy to the primary tumour and 54.25 Gy as elective dose in 33-35 fractions with or without concurrent platinum-based chemotherapy or cetuximab, whereas patients from the MDACC were treated with 66-70 Gy to the primary tumour and 56-57 Gy as elective dose in 30-33 fractions with or without concurrent platinum-based chemotherapy. In both centres, salivary glands were spared as much as possible during treatment optimisation. Additionally, starting in 2010, swallowing structures were also spared at UMCG.^21^ Refer to supplementary materials A for swallowing-sparing radiotherapy techniques.

A planning CT-scan was acquired on which the OARs were manually delineated following the guidelines of Brouwer et al. for the UMCG cohort^22^, while Atlas-based auto-contouring algorithm by Elekta ADMIRE was used for the external MDACC cohort^23^. Missing OAR segmentations were supplemented by the clinically deployed Deep Learning Contours software.^24^ Refer to supplement B for CT parameters. Dose-volume histogram parameters were extracted from the dose distributions and OAR segmentations using MATLAB (version R2018b).

### NTCP endpoint

Patients were included from the active prospective standardised follow-up programs at UMCG (NCT02435576) and MDACC (PA14-0947 data collection, PA11-0809 analysis). At UMCG, physician-rated dysphagia was scored using the adjusted Common Terminology Criteria for Adverse Events (CTCAEv4), with grade 2 (symptomatic and altered eating/swallowing) or higher at six months after radiotherapy as the NTCP endpoint. At MDACC, clinician-rated dysphagia was assessed using the “diet normalcy” question from the Performance Status Scale for Head and Neck cancer (PSS-HN), with scores of 60 or lower at 3-6 months after radiotherapy defined as the NTCP endpoint, as this cut-off corresponded best to CTCAE grade 2 dysphagia.^25^ Refer to supplement C for toxicity grading scales.

### Reference NTCP model

This study used as a reference the conventional moderate-to-severe RAD NTCP model published by Van den Bosch et al., which has been validated twice.^4,25^ The model contains the following variables: the mean dose to the oral cavity, the inferior, middle, and superior PCM, pre-treatment dysphagia score and tumour location.^4^ Updated weights for the logistic regression model were calculated in each training fold (see Supplement D), to make this NTCP model the most competitive comparator for the DL model. Performance was assessed on the independent and external test set.

### Deep learning (DL) model architecture and training

A Residual Network (ResNet) was used as DL architecture to predict RAD based on the 3D dose distribution, CT and OAR segmentations.^26^ The Tree-structured Parzen Estimator algorithm, implemented in the open-source Optuna framework, guided the fine-tuning of the model’s settings (hyperparameters).^27^ Differences in model architecture (e.g. number and size of linear layers), hyperparameters (e.g. learning rate, scheduler, batch size), and data augmentation strategies (e.g. amount of data augmentation, use of AugMix^28^) were tested. Ultimately, the optimal hyperparameters were chosen based on the mean validation loss during 5-fold cross-validation. The data augmentation and modelling were implemented using Project MONAI 0.8 in PyTorch 1.13.^29,30^ For more model training details and data preprocessing steps refer to supplementary data E.

A schematic overview of the model structure is depicted in Figure 1. The DL RAD risk prediction had two types of input: the pre-processed 3D CT-scan, 3D dose distribution and 3D OAR segmentations (hereafter called ‘3D DL-input’) and clinical variables (hereafter called ‘1D DL-input’). The 1D DL-input includes dysphagia-related variables (pre-treatment dysphagia score, pre-treatment xerostomia score, tumour location) and commonly reported patient- and treatment-related variables (T-stage, N-stage, smoking status prior to treatment, HPV-P16 status and pre-treatment WHO performance score). Multiple model configurations were tested, which are outlined in supplement F. Models were created based on only the 3D DL-input (Figure S1A: ResNet_3D_) or both the 3D and 1D DL-inputs (Figure S1B: ResNet_3D+1D(LR)_ and S1C: ResNet_3D+1D_). In ResNet_3D+1D(LR)_, two models (one based on 3D DL-input and one on 1D DL-input) are optimized separately, after which their predictions are combined using logistic regression (LR), whereas in ResNet_3D+1D_, only one model is trained and the balance between 3D and 1D DL-input is optimized simultaneously with the rest of the model.

**Figure 1.**
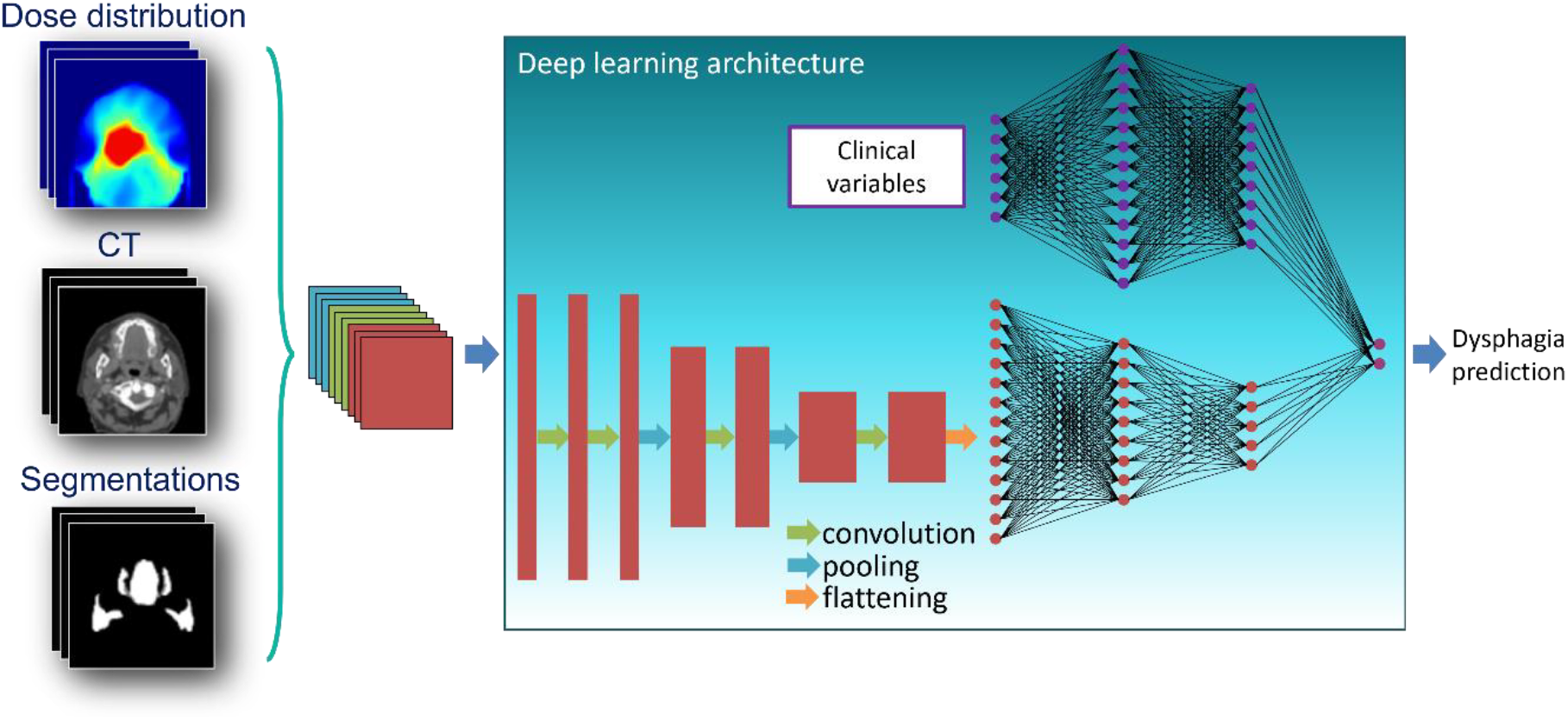
General model architecture of deep learning model. 3D DL-input of the model is the dose distribution, CT and segmentations (left) and 1D DL-input of the model is an array of patient- and treatment related (clinical) variables (middle top).

### Model evaluation

The final models were assessed using the area under the receiver-operator curve (AUC), calibration plots, Nagelkerke R^2^ and the Brier score. Metrics were calculated based on the ensemble predictions in the independent test and external test cohort, whereas the metrics were averaged over all 5 folds in the internal validation cohort. Based on these metrics, DL models were compared to the reference logistic regression NTCP model with updated weights and tested in the same cohorts.

### Leave-one-out sub-analysis

The individual contribution of the 3D DL-inputs was evaluated by performing a leave-one-out analysis on the DL NTCP models trained with all 3D DL-inputs. Instead of the true input (dose, segmentations or CT), a blank input was used to assess the impact of omitting the dose, segmentations or CT on the calibration plots.

### Attention maps

Attention maps are visual tools that highlight the most important areas in an image influencing a model’s decision. They use the model’s weights and the 3D DL-input of each individual patient to create personalised visualisations. In this study, they highlight the regions of the patient’s anatomy that contribute most to the resulting NTCP value of each individual patient. Attention maps were generated using Grad-CAM++ for the ResNet_3D_ model.^31^ Attention maps were generated in the independent test cohort for each separate cross-validation fold. They were visually evaluated to identify the regions within the head and neck area that contribute most to the prediction of RAD. Additionally, the mean attention per OAR was calculated.

## Results

### Patient characteristics

The final UMCG cohort consisted of 1112 HNC patients, which was split in 941 for the training and internal validation cohort and 171 for the independent test cohort (refer for patient exclusions to supplement G). The external MDACC test cohort consisted of 338 patients. The event rate for RAD six months after treatment were 25% in the training cohort, 26% in the independent testing cohort, and 23% in the external testing cohort. Patient characteristics did not differ between the UMCG train and test cohorts, yet were significantly different for UMCG patients and MDACC patients (Table 1).

**Table 1.**
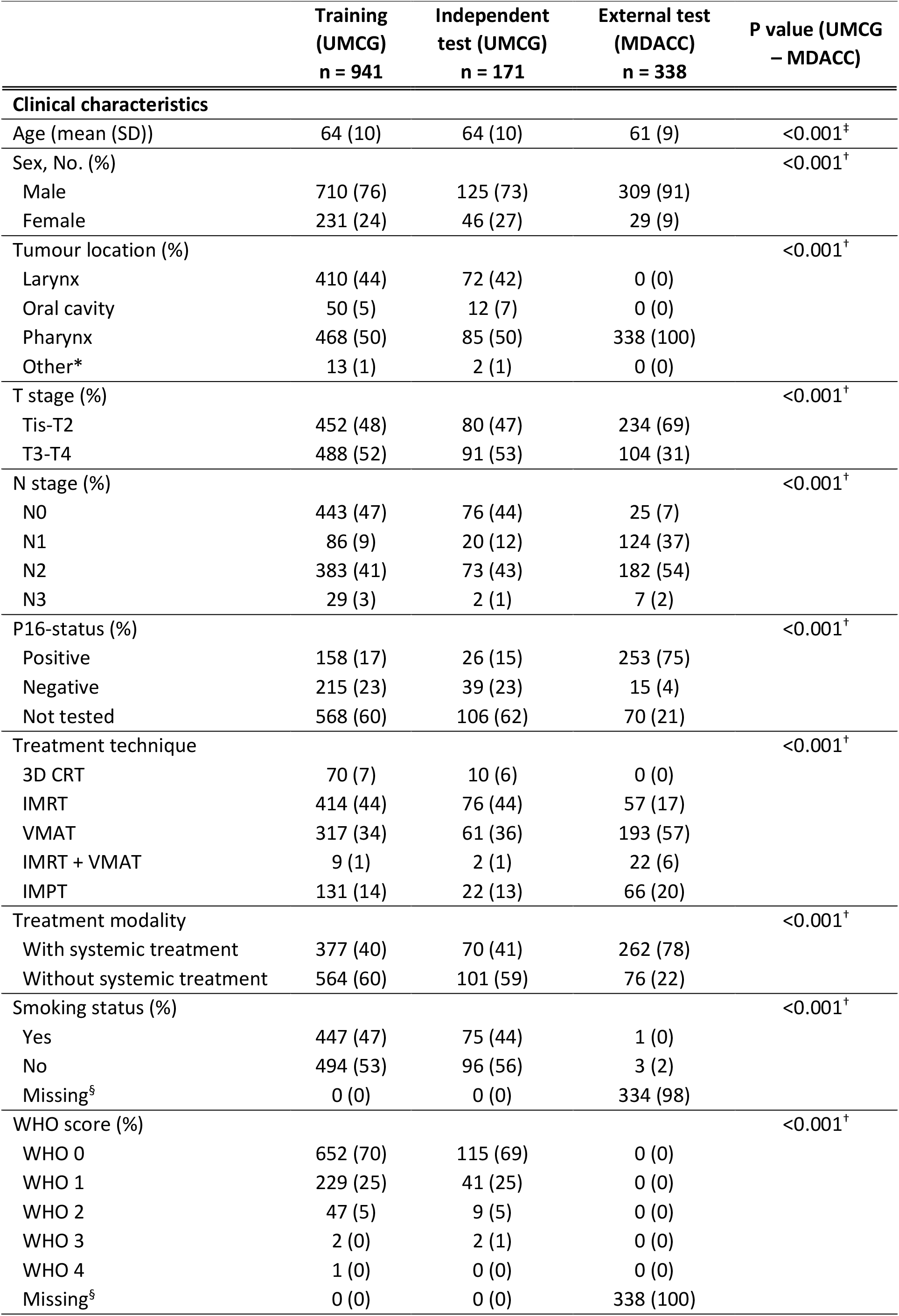
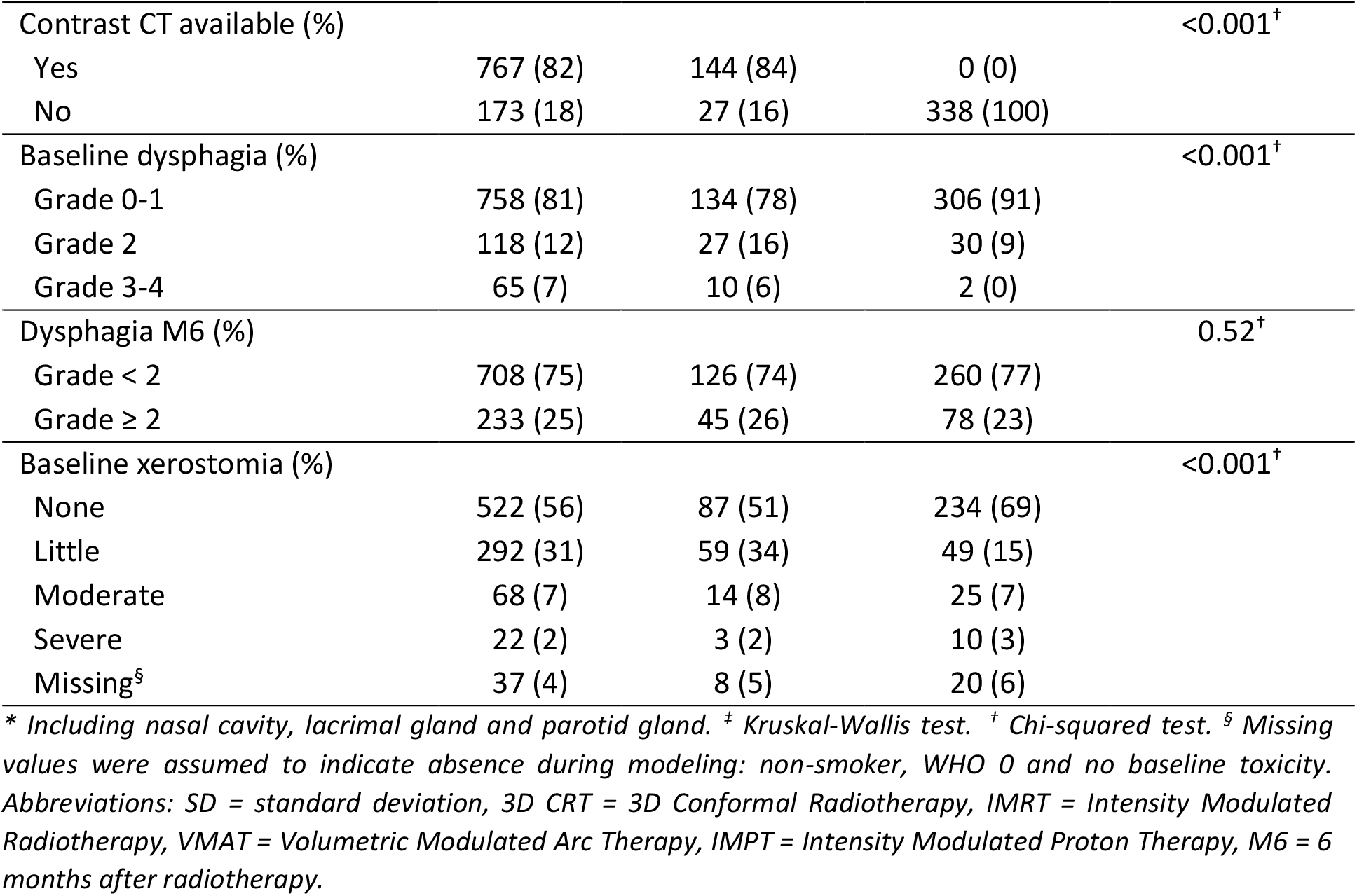
Patient characteristics.

### Model performance

The final hyperparameters that resulted in the lowest validation loss are specified per model in supplement H. All DL ResNet NTCP models outperformed the reference NTCP model across all cohorts as shown by AUC, R^2^, and Brier score (Table 2), as well as calibration (Figure 2). In the independent test cohort, the ResNet_3D_ achieved a higher AUC (AUC=0.80 [Confidence interval= 0.73-0.88]) compared to the reference model (0.76 [0.68-0.84]). The performance gain was more pronounced in the external test cohort, where the ResNet_3D_ achieved an AUC of 0.73 [0.66-0.79] versus 0.63 [0.56-0.69] for the reference model, although the overall performance decreased for all models.

**Table 2.**
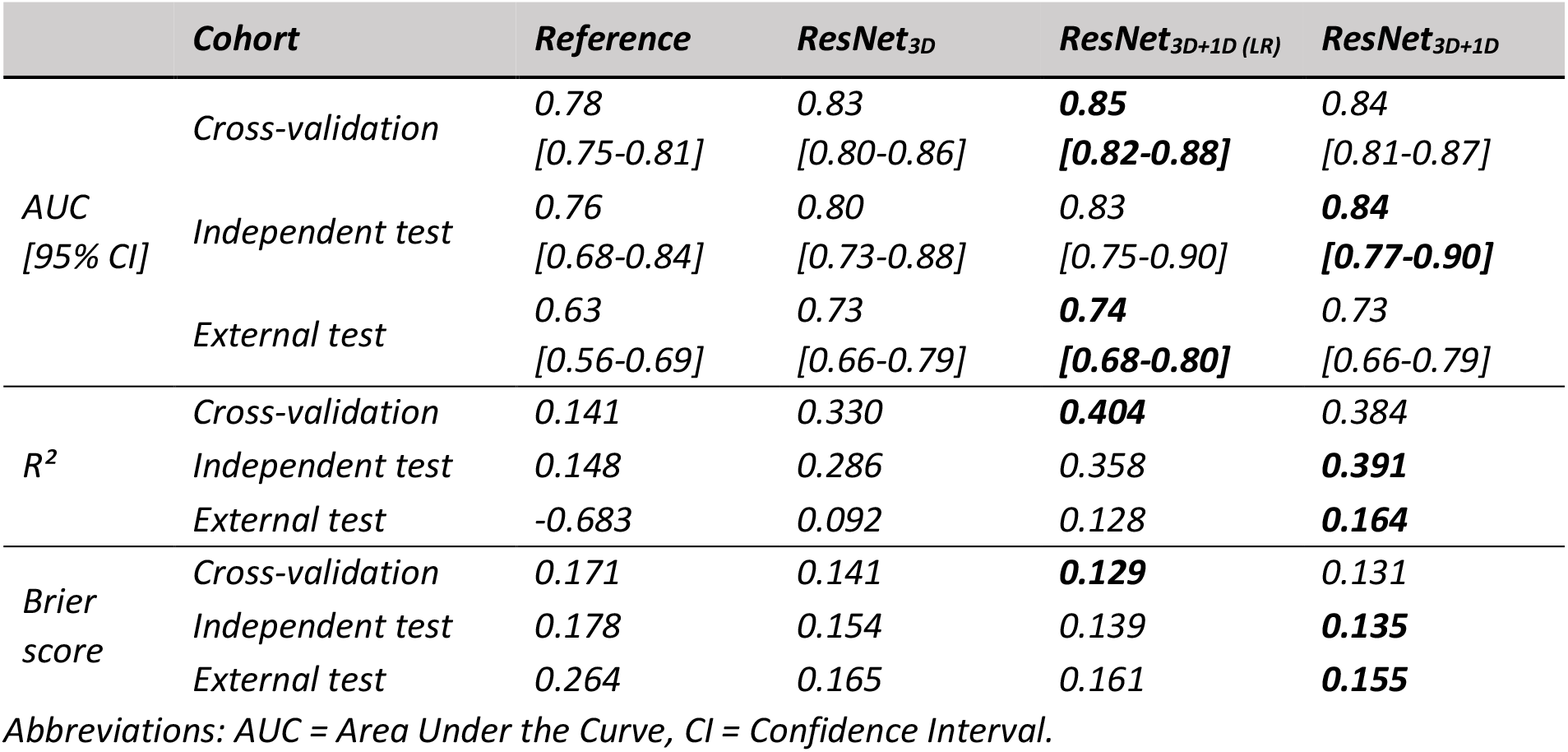
Model performance.

**Figure 2.**
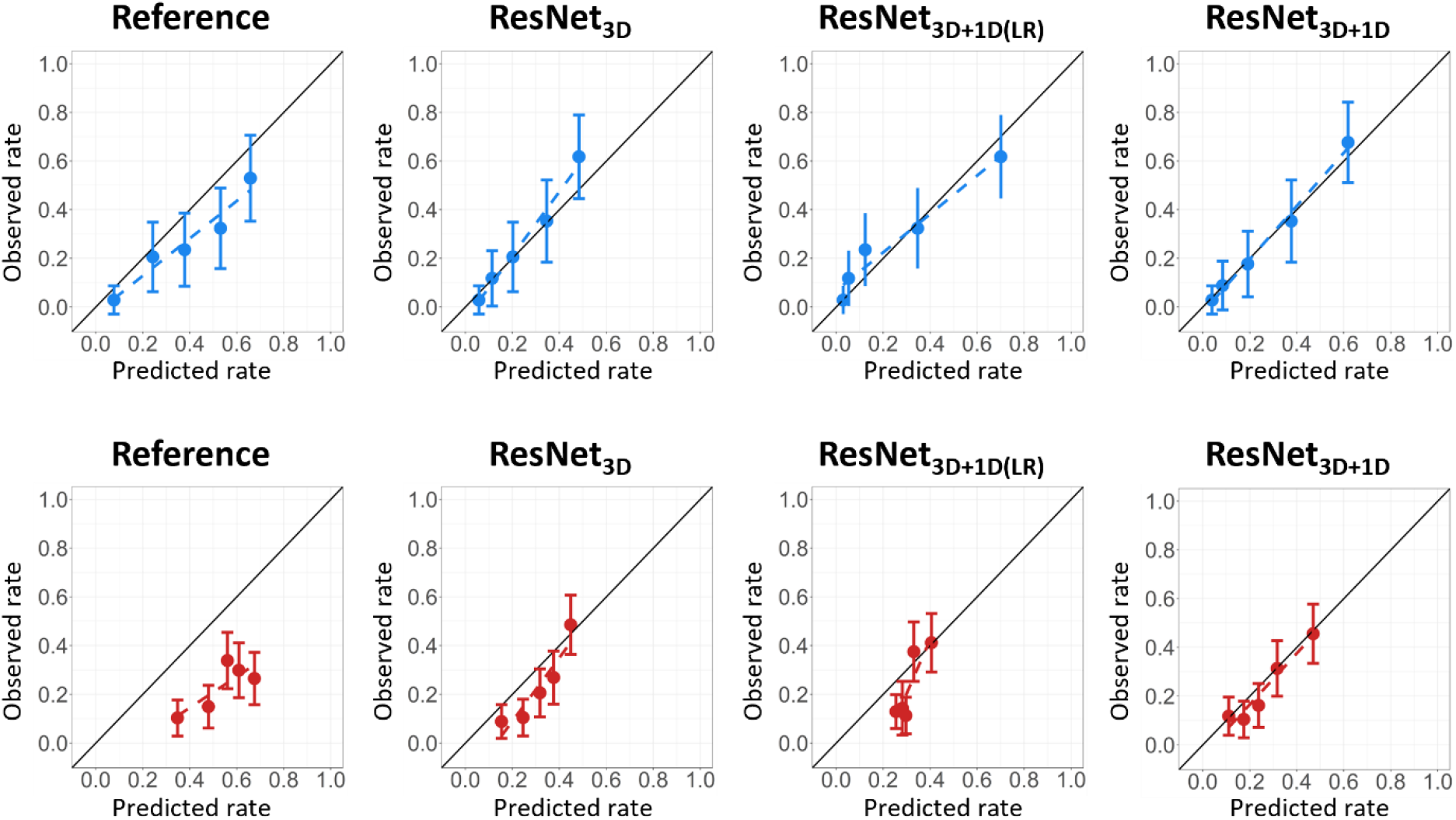
Calibration plots of model performance on independent test cohort (blue, top) and external test cohort (red, bottom).

The best overall performance was observed when incorporating both the 3D and 1D DL-inputs. In the independent test cohort, ResNet_3D+1D(LR)_ achieved an AUC of 0.83 [0.75–0.90], and the ResNet_3D+1D_ reached 0.84 [0.77–0.90]. However, the 1D DL-inputs did not improve the performance in the external test cohort, with the ResNet_3D+1D(LR)_ achieving an AUC of 0.74 [0.68–0.80] and the ResNet_3D+1D_ an AUC of 0.73 [0.66–0.79].

The R^2^ indicates that the ResNet_3D+1D_ has the best goodness-of-fit of all models with values of 0.39 and 0.16 in the independent and external test cohorts, respectively. Furthermore, this model also obtained the lowest Brier scores (0.14 and 0.16 in the independent and external test cohorts, respectively), indicating the best accuracy among these models.

Figure 2 shows that the model calibration improved in all ResNet models compared to the reference model in both the independent and external test cohort. The best calibration was achieved with ResNet_3D+1D_, demonstrating that the predicted rate closely aligns with the observed RAD rate.

### Attention maps

An attention map for a patient who developed RAD after treatment is depicted at the top of Figure 3. It shows that for this patient, the oral cavity, PCMs and salivary glands were the most important regions that contribute to the prediction of RAD at 6 months after treatment. As attention maps differ per patient, the mean attention to OARs over all patients in the independent test cohort is shown at the bottom of Figure 3. This panel shows that the oral cavity and PCM superior received the most attention across the cohort.

**Figure 3.**
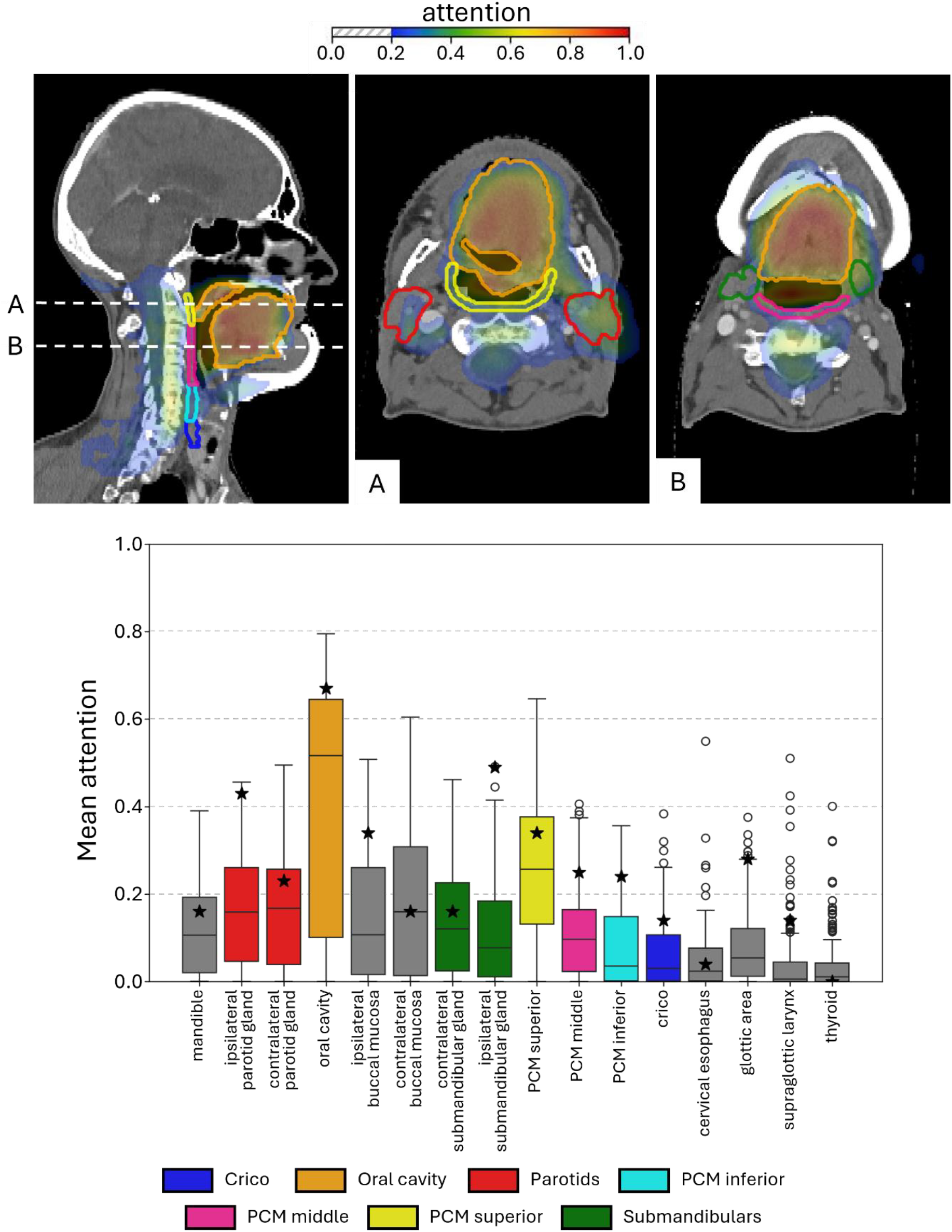
Attention maps. Top: attention map of a patient with high predicted probability for developing dysphagia, indicating that the oral cavity, PCMs, salivary glands, hyoglossus and styloglossus are regions that contribute to the prediction of dysphagia. Panel A and B are slices from height A and B. Bottom: box plot of mean attention to several OARs. The star indicates the mean attention in the OAR in the attention map of the patient at the top. Abbreviations: OAR = organ at risk, PCM = pharyngeal constrictor muscle.

### Leave-one-out sub-analysis

Omitting any input modality affected the model’s calibration performance (Figure 4). Omitting dose data led to overall lower predictions, producing a steep calibration curve. Furthermore, omitting segmentations resulted in higher predictions, and the spread in predictions became smaller for both ResNet_3D_ and ResNet_3D+1D_ (Figure 4A&C), though this effect was not observed in ResNet_3D+1D(LR)_ (Figure 4B). When CT data was omitted, predictions from ResNet_3D_ clustered around 0.5 (Figure 4A), while ResNet_3D+1D_ and ResNet_3D+1D(LR)_ showed a wider spread (Figure 4B&C).

**Figure 4.**
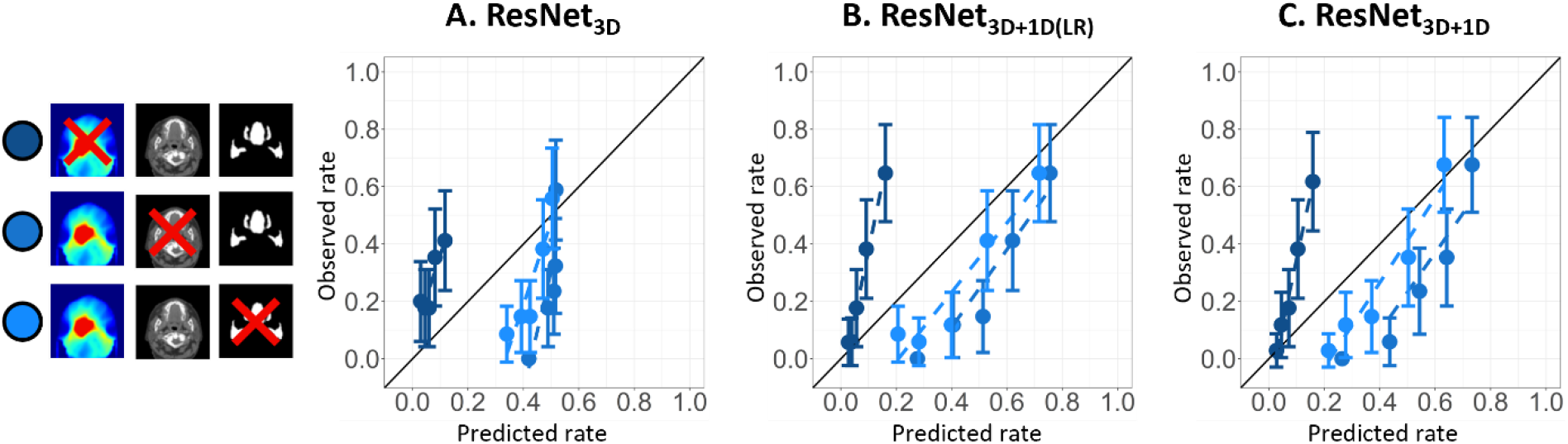
Calibration plots of leave-out-analysis in the independent test set. Darkest shade of blue: no dose, middle shade of blue: no CT, lightest shade of blue: no segmentations. Abbreviations: ResNet= Residual network.

The same analysis was performed on the external test cohort, with similar results (shown in Supplement I).

## Discussion

The 3D DL models substantially improved the prediction of RAD 6 months after radiotherapy for HNC patients compared to the 1D reference model in both an independent (AUC=0.80-0.84 vs 0.76) and external test cohort (AUC=0.73-0.74 vs 0.63). This suggests that DL models based on 3D dose distribution, CT, and segmentations capture more information regarding the development of RAD than the reference logistic regression NTCP model, which relied on 1D oral cavity and PCM substructures mean doses, tumour location and pre-treatment dysphagia score. Moreover, the addition of clinical variables improved the prediction of RAD even further in the independent test cohort (AUC=0.83-0.84 vs 0.80).

Many swallowing OARs are involved in the complex procedure to make a complete, safe and efficient swallow.^11^ As all information regarding anatomy of and dose to swallowing OARs is processed within the DL model, effects of dose on all inter-related swallowing OARs should be captured in the model. Gawryszuk et al., described the swallowing motion based on functional swallowing units (FSUs), demonstrating the importance of including more swallowing OARs in radiotherapy treatment planning and NTCP modelling for aspiration.^32^ The attention map of the individual patient (Figure 3) showed a focus of the DL network to the oral cavity, hyoglossus, and styloglossus, which are important components of the FSUs. A trend of elevated attention to the oral cavity and PCM superior was also observed in the complete independent test cohort (Figure 3, bottom).

Two methods for incorporating clinical variables in the model have been explored. In the first method, one model based on both 3D and 1D DL-input was created (ResNet_3D+1D_), while in the second method the output of two separate models (one based on 3D DL-input and one based on 1D DL-input) was combined with logistic regression (ResNet_3D+1D(LR)_). Both approaches improved performance compared to using imaging data alone (ResNet_3D_). While the differences in AUC were limited, the direct integration (ResNet_3D+1D_) showed more consistent calibration and better goodness-of-fit, particularly in the external test cohort. This aligns with expectations, as during model training, model weights for the parts processing 3D DL-input and 1D DL-input are optimized simultaneously, directly balancing all information from both DL-input types. In ResNet_3D+1D(LR)_, this balance is achieved by combining two separate models, meaning the 3D DL-input and 1D DL-input are processed independently. This prevents the model from establishing a specific relationship between, e.g. a dose parameter from the 3D DL-input and the pre-treatment dysphagia score from the 1D DL-input, which would be possible in ResNet_3D+1D_.

In the leave-one-out-analysis, omitting radiation dose from the 3D DL-input resulted in lower predictions compared to when all input was available (Figure 4), aligning with the intuitive reasoning that no dose implies no expected toxicity. Omitting CT or segmentations from the 3D DL input caused risk overestimation in both test cohorts (Figure 4&S4), suggesting the model uses them to lower RAD predictions. Overall, this suggests that the DL models heavily rely on dose to swallowing OARs.

As post-treatment RAD is related to damage to swallowing muscles, an insight into the pre-treatment structural status of these muscles may provide more information for the prediction of RAD.^11^ In our modelling approach, CTs were used to incorporate information about anatomy, but muscle mass, and especially muscle quality, can be better determined by MRI than by CT, due to the improved soft tissue contrast.^33^ Pre-treatment sarcopenia, a type of muscle loss that can be assessed using MRI, is related to the development of RAD after treatment.^34^ Therefore, developing a model that contains MRI in the 3D DL-input would be promising to improve the prediction of RAD even further.

One of the main limitations of this study was that there were significant differences in patient characteristics between the UMCG and MDACC cohorts, as well as the endpoint assessment between centres (CTCAE vs PSS-HN diet normalcy score), potentially influencing results in the external test cohort. This could explain why the performance measures were lower for both the reference and DL models in the external test. Nevertheless, the external results still showed a large performance increase of the DL models compared to the 1D reference NTCP model. Additionally, the (sub)components of the FSUs were not delineated per guideline^35^, as this approach is highly time-consuming, making it impractical for 1,484 patients. Consequently, they were not included as segmentations in the DL models, and the attention map analysis in the independent test set was limited to 16 OARs from the guidelines by Brouwer et al.^22^ Furthermore, dysphagia assessment in this study relied on clinician-graded measures that essentially reflect restrictions in oral intake. Oral intake is a notoriously non-specific measure of function. In HNC, impaired oral intake reflects oral and pharyngeal dysfunction, but also salivary, taste, appetite, and pain symptoms. As such, more comprehensive and swallowing specific imaging-based assessments such as Modified Barium Swallowing studies (MBS) are expected to yield more precise RAD-specific models.^36^ MBS has been used in previous studies to assess how swallowing patterns are altered by radiation dose, which could offer a more accurate prediction of RAD based on radiation dose.^37,38^ In a future study, our 3D modelling approach could be combined with MBS-based outcome metrics, e.g. the DIGEST score^39^, to improve the prediction of RAD by finding stronger relations between the affected swallowing structures and the dose that was delivered. Lastly, the size of our development cohort may be insufficient given the complexity of the DL model, as large, high-quality datasets are essential for robust model training and these cohorts are limited in the medical domain. Developing a DL model on larger multi-centre datasets could provide a solution to this problem.

## Conclusion

The 3D DL NTCP models predicting RAD after radiotherapy outperformed the reference 1D NTCP model in both the independent and the external test cohort based on discrimination and calibration. Adding clinical variables to the 3D DL NTCP model improved the performance even further. These results suggest that RAD prediction can be improved by using the entire 3D dose distribution, OAR segmentations, the CT scan and clinical variables.

## Data Availability

All data produced in the present study are available upon reasonable request to the authors.

## Acknowledgements

We would like to thank the Center for Information Technology of the University of Groningen for their support and for providing access to the Hábrok high performance computing cluster. Collection of the external data from MDACC was realised by the generous support by Mr. and Mrs. Charles W. Stiefel at The University of Texas MD Anderson Cancer Centre-Oropharynx Cancer Program and currently maintained by infrastructure support via National Cancer Institute Award 1P01CA285249.

