## Supplementary data for "Deep learning NTCP model for late dysphagia after radiotherapy for head and neck cancer patients based on 3D dose, CT and segmentations"

### A. Swallowing-sparing radiotherapy at UMCG

Swallowing-sparing radiotherapy has been implemented at the department of Radiotherapy at the UMCG since 2010, utilizing normal tissue complication probability (NTCP) models. The table below summarizes the organs at risk (OARs) that have been selectively spared using various NTCP models.

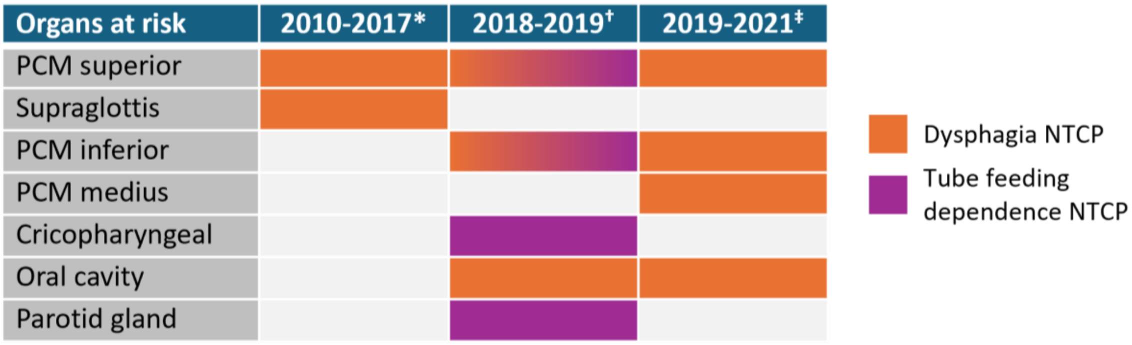

*Model by Christianen et al. (2012) [1]

† Model by Christianen et al. (2012) and model by Wopken et al. (2014) [2], updated by Langendijk et al. (2021) [3]

‡ Model by Van den Bosch et al. (2021) [4], updated for clinical usage in the National Indication Protocol Protons version 2.2

NTCP models follow the following equation:

$$NTCP = \frac{1}{1 + e^{-S}}$$

#### Model 2010 - 2017

$$S_{dysphagia} = -6.09 + 0.057 \times D_{mean}\ \text{PCM superior} + 0.037 \times D_{mean}\ \text{Supraglottis}$$

#### Models 2018 - 2019

$$\begin{aligned} S_{dysphagia} = & -3.303 + 0.024 \times D_{mean}\ oral\ cavity \\ & + 0.024 \times D_{mean}\ PCM\ superior \\ & + 0.967 \times baseline\ dysphagia\ (1\ if\ baseline\ dysphagia\ is\ grade\ \geq 2, 0\ if\ not) \end{aligned}$$

$$\begin{aligned} S_{tube\ feeding\ dependence} = & -6.849 + 0.680 \times advanced\ T\ stage\ (1\ if\ T3\ or\ T4, 0\ if\ not) \\ & + 0.317 \times moderate\ weigth\ loss\ (1\ if\ 1 - 10\%, 0\ if\ not) \\ & + 1.178 \times severe\ weight\ loss\ (1\ if\ > 10\%, 0\ if\ not) \\ & + 0.198 \times accelerated\ radiotherapy\ (1\ if\ accelerated\ RT, 0\ if\ not) \\ & + 1.101 \times chemoradiation\ (1\ if\ chemoradiation, 0\ if\ not) \\ & + 1.716 \times radiotherapy\ plus\ cetuximab\ (1\ if\ RT\ and\ cetuximab, 0\ if\ not) \\ & + 0.030 \times D_{mean}\ PCM\ superior \\ & + 0.013 \times D_{mean}\ PCM\ inferior \\ & + 0.022 \times D_{mean}\ contralateral\ parotid\ gland \\ & + 0.008 \times D_{mean}\ cricopharyngeal\ muscle \end{aligned}$$

#### Model 2019 - 2021

$$\begin{aligned} S_{dysphagia} = & -4.0536 + 0.0300 \times D_{mean}\ oral\ cavity \\ & + 0.0236 \times D_{mean}\ PCM\ superior \\ & + 0.0095 \times D_{mean}\ PCM\ medius \\ & + 0.0133 \times D_{mean}\ PCM\ inferior \\ & + 0.9382 \times baseline\ dysphagia\ (1\ if\ baseline\ dysphagia\ is\ grade\ 2, 0\ if\ not) \\ & + 1.2900 \times baseline\ dysphagia\ (1\ if\ baseline\ dysphagia\ is\ \geq grade\ 3, 0\ if\ not) \\ & - 0.6281 \times tumour\ location\ (1\ if\ tumor\ location\ is\ pharynx, 0\ if\ not) \\ & - 0.7711 \times tumour\ location\ (1\ if\ tumor\ location\ is\ larynx, 0\ if\ not) \end{aligned}$$

#### B. CT scanner parameters

Table S1: CT scanner parameters.

| Manufacturer & model name | Kilo voltage peak [kV] | Slice thickness [mm] | Spacing [mm] | Years in use | n patients |
|---|---|---|---|---|---|
| SIEMENS Sensation Open | 120 | 2-4 | 0.85-1.60 | 2007-2015 | 356 |
| SIEMENS Biograph 64 | 80-120 | 2 | 0.56-1.52 | 2010-2019 | 378 |
| SIEMENS SOMATOM Definition AS | 100-140 | 2 | 0.90-1.56 | 2015-2021 | 372 |
| Siemens Healthineers SOMATOM go.Open Pro | 100-120 | 2 | 0.98-1.56 | 2020-2021 | 6 |
| **Median (IQR)** | 120 (100-120) | 2 (2-2) | 0.98 (0.98-0.98) | - | - |

*Abbreviations:* IQR = interquartile range.

#### C. Toxicity assessment scales

Table S2 Adjusted Common Terminology Criteria for Adverse Events (CTCAE) version 4 dysphagia. The NTCP endpoint in this study was grade 2 or higher dysphagia.

| SCORE | DESCRIPTION |
|---|---|
| **0** | Asymptomatic, able to eat regular diet |
| **1** | Symptomatic, able to eat regular diet |
| **2** | Symptomatic and altered eating/swallowing |
| **3** | Severely altered eating/swallowing: tube feeding or TPN or hospitalization indicated |
| **4** | Life-threatening consequences; urgent intervention indicated |
| **5** | Death |

Table S3 Performance Status Scale for Head and Neck cancer (PSS-HN) diet normalcy. The NTCP endpoint in this study was a PSS-HN score of 60 or lower.

| SCORE | DESCRIPTION |
|---|---|
| **100** | Full diet (no restrictions) |
| **90** | Full diet (liquid assist) |
| **80** | All meat |
| **70** | Raw carrots, celery |
| **60** | Dry bread and crackers |
| **50** | Soft chewable foods (e.g. macaroni, canned/soft fruits, cooked vegetables, fish, hamburger, small pieces of meat) |
| **40** | Soft foods requiring no chewing (e.g. mashed potatoes, apple sauce, pudding) |
| **30** | Pureed foods (in blender) |
| **20** | Warm liquids |
| **10** | Cold liquids |
| **0** | Non-oral feeding (tube fed) |

#### D. Updated weights logistic regression model

A refitting of the reference dysphagia NTCP model by Van den Bosch et al. was required, as patients from its development cohort are also included in the independent test cohort of the deep learning model developed in this study.[4] Using the original model without adjustment could lead to an overestimation of performance due to this cohort overlap. Hence, in each of the 5 training folds, new coefficients are estimated based on the training data.

The reference NTCP model was developed using a method that addresses multicollinearity by grouping predictors such that each group contains predictors with low correlation between each other. Logistic regression NTCP models are then made with each predictor group, called sub-models. These sub-models are combined into a single composite model, representing the final NTCP model.[5]

For the grade ≥2 dysphagia model, these are the predictors in the sub-models:

*(s1)* $D_{mean}$ Oral cavity (OC), $D_{mean}$ PCM middle (PCMm), $D_{mean}$ PCM inferior (PCMi), Baseline toxicity (BSL), Tumour location (TL)

*(s2)* $D_{mean}$ PCM superior (PCMs), $D_{mean}$ PCM middle (PCMm), $D_{mean}$ PCM inferior (PCMi), Baseline toxicity (BSL), Tumour location (TL)

To construct the composite model, the coefficients from sub-models 1 and 2 are averaged. The final NTCP formula thus becomes:

$$NTCP = \frac{1}{1 + e^{-L}}, with$$

$$\begin{aligned} L = & \frac{Intercept_{s1} + Intercept_{s2}}{2} \\ & + \beta_{s1,OC} \cdot D_{mean}\ OC \\ & + \beta_{s2,PCMs} \cdot D_{mean}\ PCMs \\ & + \frac{\beta_{s1,PCMm} + \beta_{s2,PCMm}}{2} \cdot D_{mean}\ PCMm \\ & + \frac{\beta_{s1,PCMi} + \beta_{s2,PCMi}}{2} \cdot D_{mean}\ PCMi \\ & + \frac{\beta_{s1,BSL\ grade\ 2} + \beta_{s2,BSL\ grade\ 2}}{2} \cdot BSL\ grade\ 2 \\ & + \frac{\beta_{s1,BSL\ grade\ 3-4} + \beta_{s2,BSL\ grade\ 3-4}}{2} \cdot BSL\ grade\ 3-4 \\ & + \frac{\beta_{s1,TL\ pharynx} + \beta_{s2,TL\ pharynx}}{2} \cdot TL\ pharynx \\ & + \frac{\beta_{s1,TL\ larynx} + \beta_{s2,TL\ larynx}}{2} \cdot TL\ larynx \end{aligned}$$

With $\beta_{s1}$ the coefficient of sub-model 1 and $\beta_{s2}$ the coefficient of sub-model 2. This approach was applied within each of the 5 training folds. For each fold, an NTCP value was determined per patient using the corresponding coefficients for the model parameters. The metrics of the cross-validation are based on averaged predictions, whereas the metrics of the independent and external test cohorts are determined by averaging the metric over the folds, similar to the deep learning NTCP values.

### E. Data preprocessing & (Optuna) hyperparameter optimization

#### 3D data preprocessing

For each patient, the 3D CT-scan, 3D dose distribution and OAR segmentations were resampled to voxels of 2x2x2 $mm^3$ (median original voxel sizes CT: 0.98x0.98x2 $mm^3$). Subsequently, all input images were cropped to a bounding box between the thyroid contour (lower boundary) and the top of the parotid glands contours (upper boundary). The centre of this bounding box was used as the centre of the final volumes of 100x100x100 voxels. The CT values, in Hounsfield Units (HU), were clipped between -200 and 400 HU and the dose distribution was clipped between 0 and 8000 cGy. The CT and dose data were normalised between 0 and 1. The following binary mask OARs were used as input for the dysphagia DL model: oral cavity, PCMs, parotid glands, submandibular glands, and cricopharyngeal muscle. Refer to Chu et al. (2024) for more detailed preprocessing steps.[6]

#### Set parameters

Table S4 Parameters that were always kept constant during hyperparameter optimization.

| PARAMETER | SET VALUE |
|---|---|
| **Weight of labels segmentations** | |
| *Pharyngeal constrictor muscles* | 1 |
| *Oral cavity* | 1 |
| *Cricopharyngeal muscle* | 0.5 |
| *Parotid glands* | 0.5 |
| *Submandibular glands* | 0.5 |
| **Leaky ReLU alpha** | 0.1 |
| **Weight initialization** | Kaiming uniform |
| **Early stopping** | After 10 epochs no validation AUC increase |
| **Loss function** | Cross-entropy |
| **Starting learning rate** | 0.0001 |

#### Suggested hyperparameters Optuna

Table S5 The hyperparameters that could be suggested per trial to the Optuna hyperparameter optimization framework.[7]

| PARAMETER | OPTIONS |
|---|---|
| **Batch size** | 2, 4, 8, 16 |
| **Amount linear layers** | 1, 2, 3 |
| **Amount of nodes per linear layer** | 8, 16, 32, 64 |
| **Dropout chance in linear layers** | Between 0 and 0.5 |
| **Amount linear layers for clinical variables*** | 1, 2, 3 |
| **Amount of nodes per linear layer for clinical variables*** | 8, 16, 32, 64 |
| **Dropout chance in linear layers for clinical variables*** | Between 0 and 0.5 |
| **Linear layer in which the clinical variables and the ResNet output will be combined*** | Between 0 (first layer after global average pooling layer) and the amount of linear layers chosen by Optuna minus 1 |
| **Data augmentation strength** | Between 0 and 3 |
| **Data augmentation probability** | Between 0 and 1 |
| **Perform AugMix** [8] | True[†] or False |
| **AugMix strength** | Between 0 and 3 |
| **Optimizer name** | AdaBelief[9] or MADGRAD[10] |
| **Scheduler** | Cosine or cyclic |

| | |
|---|---|
| **T0 (cosine scheduler)** | Between 8 and 64 |
| **Step_size_up (cyclic scheduler)** | 96* batch size or 64* batch size or 32*batch size |
| **Use momentum** | True or False |
| **Momentum (if optimizer = MADGRAD)** | Between 0.8 and 1 |
| **Label smoothing** | Between 0 and 0.1 |
| **Use bias** | True or False |

* Only applicable for models including 1D DL-input, † If AugMix was performed, the mixture width was 3 by default and mixture depth was [1,3] by default.

#### Optuna settings

The Tree-structured Parzen Estimator algorithm, implemented in the open-source Optuna framework, was applied for 100 trials to find the optimal hyperparameters from the search space. The objective was to minimize the average validation loss over 5-fold cross-validation. The version of Optuna that was used was 3.3.0.

#### Multi-layer perceptron grid search

The multi-layer perceptron was not optimized using Optuna, but by using a grid-search of optimal hyperparameters. All combinations that can be made using the parameters in the table below were tested. The multi-layer perceptron was trained independently from the part of the network which includes the CT, dose distribution and segmentations. The configuration of the parameters with the lowest validation loss was used to make a combined model of clinical variables and imaging/dose characteristics.

Table S6 Hyperparameters multi-layer perceptron.

| PARAMETER | OPTIONS |
|---|---|
| **Fully connected layers** | 1, 2, 3 |
| **Nodes per layer** | 8, 16, 32, 64 (same size or smaller in next layer) |
| **Dropout probability** | 0, 0.1, 0.2, 0.3, 0.4, 0.5 |

#### Data augmentation

Two methods of data augmentation have been explored. The AugMix algorithm and standard augmentation using flipping, translating, zooming, intensity shifting and rotation.[8] If both are used during training, first standard data augmentation was applied, followed by AugMix. Which data augmentation steps are used is optimized using Optuna.

Standard data augmentation possibilities:

- Flipping*: yes/no, only in transversal plane
- Translating*: translating 7 times the data augmentation strength in pixels
- Zooming*: zooming with a factor of 0.07 times the data augmentation strength + 1 in every direction.
- Intensity shifting: offsets is between 0.05 times the data augmentation strength and minus 0.05 times the data augmentation strength. The intensity in the whole image will be shifted with this value.
- Rotating*: rotating the image in the transversal plane with 12° times the data augmentation strength

Augmentations that are also incorporated in AugMix are indicated with an * in the list above.

Regardless of whether other data augmentation steps were used, the 3D inputs were always cropped to a size of 96x96x96 pixels.

#### F. Detailed model architecture

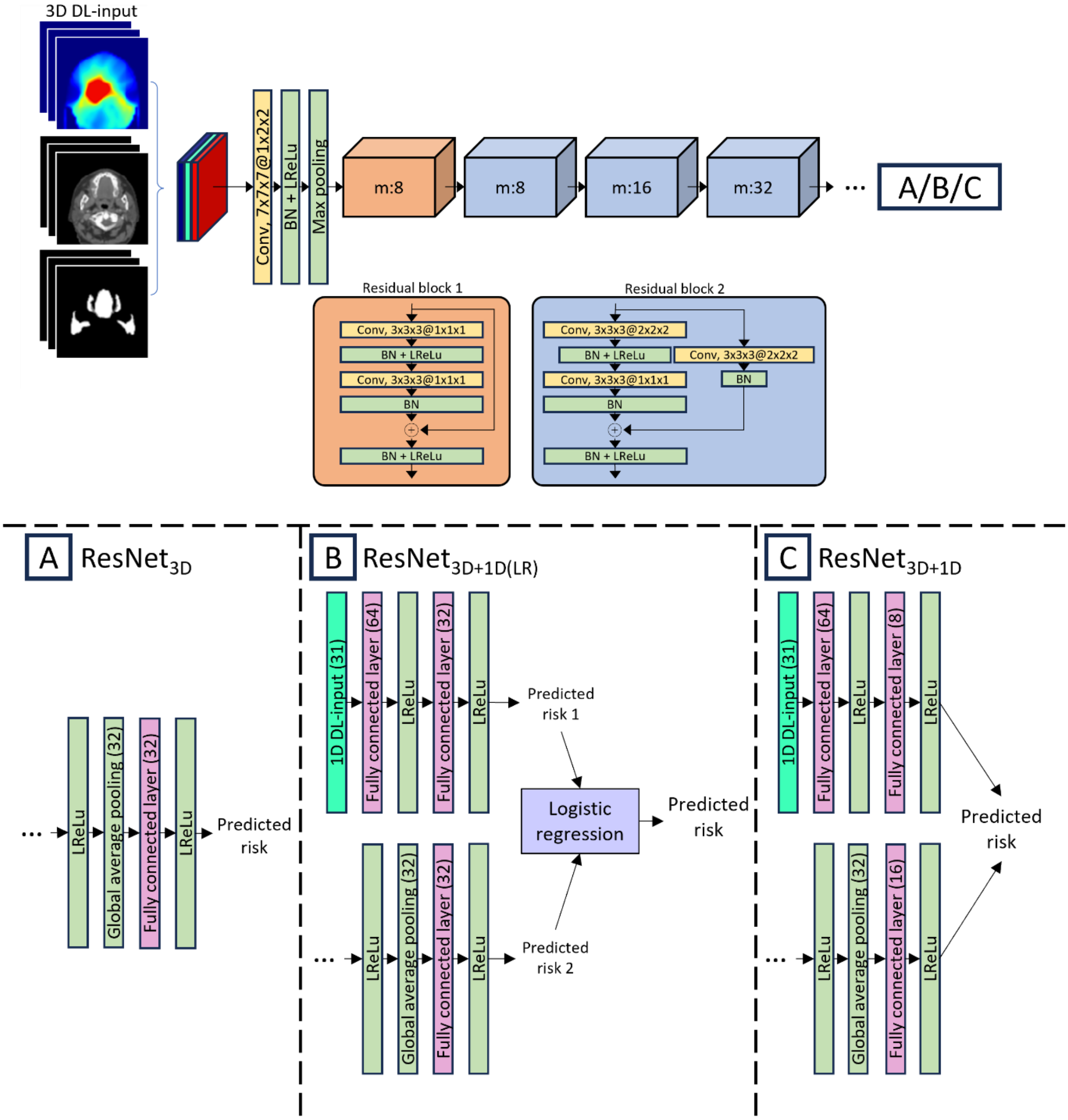

Figure S1 Detailed model architecture. The body of the model, a ResNet, is depicted at the top. Three different heads of the model are depicted on the bottom, represented by A, B and C. The heads are added to the ResNet after the first Leaky ReLu layer after the last residual block. A. Model architecture without 1D DL-input. B. Adding a separate multi-layer perceptron to the model architecture. The predictions of the model based on 3D DL-input and the model based on 1D DL-input are combined using logistic regression. The model based on 3D DL-input is the ResNet$_{3D}$ of image S1A. C. Adding clinical variables by incorporating fully connected layers. The model processed 1D and 3D inputs in parallel through separate branches, which were merged within the network to produce a unified prediction. *Abbreviations:* 3D DL-input = dose + segmentations + CT, 1D DL-input = clinical variables, Conv = convolutional layer, BN = Batch Normalization, LReLu = Leaky Rectified Linear Unit, m = output channels.

#### G. Exclusions

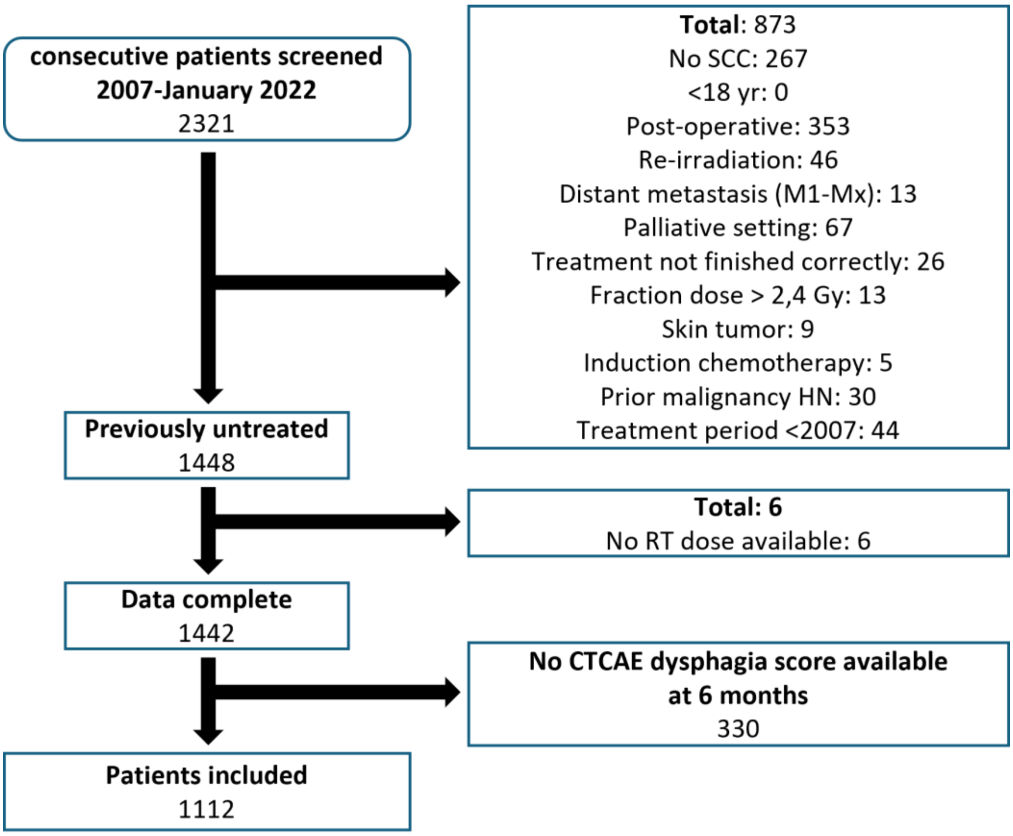

Figure S2 Exclusions for the UMCG cohort.

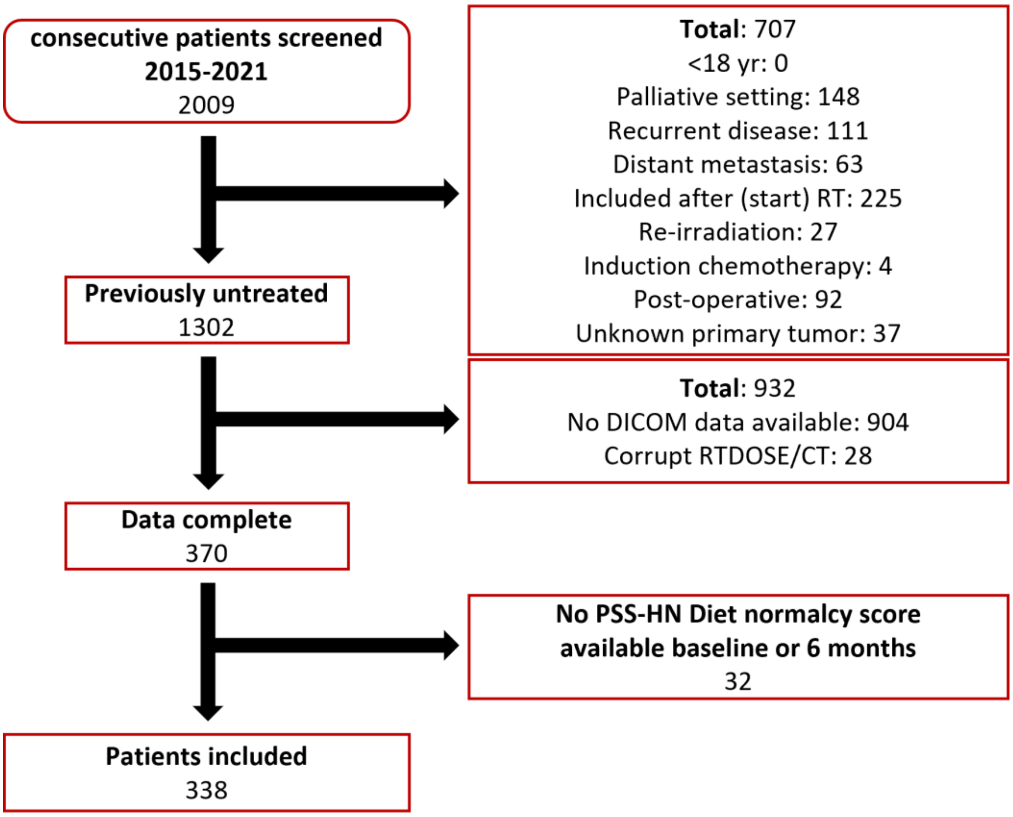

Figure S3 Exclusions for the MDACC cohort.

#### H. Results hyperparameter optimization

##### Optimal hyperparameters Optuna

In the table below, the combinations of hyperparameters that gave the lowest validation loss for the deep learning models with and without 1D DL-input are displayed.

Table S7. Final hyperparameters

| PARAMETER | RESNET$_{3D}$ | RESNET$_{3D+1D}$ |
|---|---|---|
| **Batch size** | 4 | 4 |
| **Amount linear layers** | 1 | 1 |
| **Amount of nodes per linear layer** | 32 | 16 |
| **Dropout chance in linear layers** | 0.45 | 0 |
| **Amount linear layers for clinical variables** | - | 2 |
| **Amount of nodes per linear layer for clinical variables** | - | 64, 8 |
| **Dropout probability in linear layers for clinical variables** | - | 0.15, 0.50 |
| **Linear layer in which the clinical variables and the ResNet output will be combined** | - | 0 (end) |
| **Data augmentation strength** | 0.5 | 0.55 |
| **Data augmentation probability** | 0.7 | 0.3 |
| **Perform AugMix** [8] | True | False |
| **AugMix strength** | 0.2 | - |
| **Optimizer name** | AdaBelief [9] | MADGRAD [10] |
| **Scheduler** | Cyclic | Cyclic |
| **T0 (cosine scheduler)** | - | - |
| **Step_size_up (cyclic scheduler)** | 64 * batch size | 64 * batch size |
| **Use momentum (if optimizer = MADGRAD)** | False | False |
| **Momentum (if optimizer = MADGRAD)** | - | - |
| **Label smoothing** | 0.02 | 0 |
| **Use bias** | False | True |

##### Multi-layer perceptron grid search

The hyperparameters with the lowest validation loss are listed in the table below.

Table S8. Final hyperparameters grid search.

| PARAMETER | VALUE |
|---|---|
| **Fully connected layers** | 2 |
| **Nodes per layer** | 64, 32 |
| **Dropout probability** | 0.2, 0.2 |

#### Logistic regression for combination of deep learning model predictions and multi-layer perceptron predictions

To combine the $ResNet_{3D}$ predictions with the multi-layer perceptron predictions, a logistic regression model was trained based on the cross-validation set. The coefficients from this model are listed in the table below.

Table S9. Final logistic regression coefficients for $ResNet_{3D+1D(LR)}$

| PARAMETER | COEFFICIENTS |
|---|---|
| **Intercept** | -4.464 |
| **Deep learning prediction** | 7.247 |
| **Multi-layer perceptron prediction** | 4.888 |

#### I. Calibration plots leave-out analysis external cohort

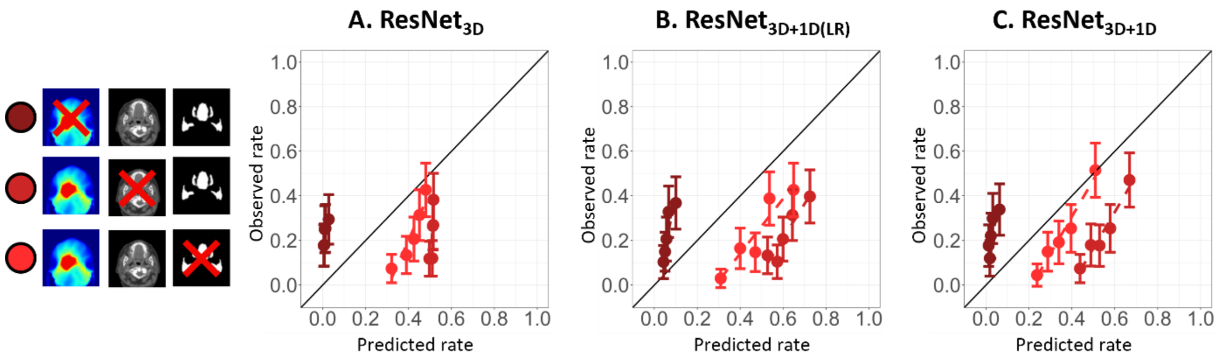

Figure S4. Calibration plots leave-out analysis external test cohort. Darkest shade of red: no dose, middle shade of red: no CT, lightest shade of red: no segmentations. Abbreviations: ResNet= Residual network.
